# Development and implementation of an AI system for clinical toxicology sign-outs

**DOI:** 10.64898/2026.01.29.26345133

**Authors:** Nathan Laha, Michael Keebaugh, Hsuan-Chieh Liao, Bright Amankwaa, Olumuyiwa Adesoye, Abed Pablo, William S Phipps, Andrew N Hoofnagle, Geoffrey S Baird, Patrick C Mathias, Brody H Foy

## Abstract

**Background:** Modern natural language tools have potential to improve clinical workflows, but few have been successfully deployed in practice. Here, we present the development, deployment, and evaluation of an AI language tool for generating preliminary clinical sign-outs in a urine drug testing service.

**Methods:** Large language models (LLMs) were used to extract substance use patterns from 83,553 urine drug test interpretations. We then trained an AI model using these data to predict substance use from qualitative and quantitative urine testing results. Predicted substance use patterns were used to create preliminary clinical sign-out statements, which were then integrated into an existing clinical workflow. Pre- and post-deployment user studies were performed to evaluate model performance and user experience within this workflow.

**Results:** LLM-based extraction of substance-use patterns was 99.9% accurate, outperforming human labelling. Substance use prediction was similarly accurate, with area under the ROC curve > 0.99 across 33 drug categories. Workflow integration reduced clinical sign-out times by 65s per case (51% efficiency gain), with the greatest benefits seen for less experienced users.

**Conclusions:** AI-based interpretation of urine drug testing was fast and accurate, providing significant efficiency gains to the clinical service. This demonstrates that natural language tool integration can provide substantial clinical benefit, without comprising quality of care.

## Introduction

Urine drug tests (UDTs) are employed in diverse clinical care settings, including emergency medicine, chronic pain management, and addiction medicine^1–3^. For patients with substance-use disorders in particular, UDTs provide objective data to monitor medication adherence and support treatment adjustments^3,4^.

Many UDT test results and interpretations are straightforward; however, some cases involve complex pharmacokinetics, overlapping metabolic pathways, or assay cross-reactivity^2,5^. In these cases, ordering providers may benefit from consultation with a clinical chemist with toxicology expertise, to assess the most likely substance use pattern^6^. Although valuable, such consultations can be time intensive, leading to increased turnaround times and operational costs.

Recent advances in natural language processing (NLP) tools, particularly large-language models (LLMs) offer potential to reduce these bottlenecks^7,8^. Modern LLMs have the ability to extract and process multi-modal clinical data to produce textual outputs^7,9^. As such, there is wide interest in use of LLMs to enhance clinical sign-outs across medicine^9^. Yet clinical deployment of LLMs remains rare, due in large part to the risks that generative models pose – such as hallucinations or generating outputs that deviate from regulatory compliance rules^10–13^.

Within this context, we investigated whether machine learning (ML) tools could enhance toxicology service efficiency. We used LLMs to develop a large structured dataset from historical toxicology sign-out notes, which was then used to train a classical ML model for substance use prediction. Resultant predictions were used to generate AI-based clinical sign-out notes, which were incorporated into a human-in-the-loop sign-out workflow. This approach achieved expert-level accuracy while significant reducing sign-out time, demonstrating a practical framework for integrating AI and NLP tools into complex clinical workflows.

## Methods

### Urine drug testing workflow

At UW Medicine (UWM), UDTs ordered for monitoring of controlled substance use followed a two-stage process^6^. Samples were first evaluated using qualitative immunoassay screening, which detected the use of amphetamines, barbiturates, benzodiazepines, buprenorphine, cannabinoids, cocaine, fentanyl, methadone, opiates, oxycodone, and tramadol. When indicated, additional confirmatory testing was performed via mass spectrometry, which detected amphetamines (amphetamine, 3-4- methylenedioxyamphetamine [MDA], 3-4-methylenedioxymethamphetamine [MDMA], phentermine, and ritalinic acid), benzodiazepines (bromazolam, estazolam, flualprazolam, flubromazolam, lorazepam, nordiazepam, oxazepam, temazepam, and metabolites of: clonazepam, flurazepam, flunitrazepam, midazolam, triazolam, flurazepam, zolpidem, and zopiclone), and opiates (buprenorphine, codeine, fentanyl, hydrocodone, hydromorphone, meperidine, methadone, morphine, oxycodone, oxymorphone, propoxyphene, tramadol, and associated metabolites). Alongside these tests, an enzymatic reaction was used to detect ethanol (alcohol). The results were first reviewed and interpreted by a clinical chemistry reader (a pathology resident, fellow, or clinical toxicologist), and were then signed out by a board-certified clinical pathologist (WSP, ANH, or GSB). From Feb-2021 onwards, case sign-outs were performed using an advanced web application interface, developed by UWM to support this service^14,15^.

### Clinical data

We collated all UWM UDT test results with interpretative notes from Jan-01-2014 to Feb-29-2024 (n: 83,553). Data was pulled via SQL queries to UWM’s health record databases. The set of detectable substances varied over this time period due to changes in UWM’s patient population and laboratory testing protocols. For this study, we restricted analysis to all substances that were detectable under 2024 protocols, and had a detected prevalence above 0.1% (23 substances total). These substances were: alcohol (ALC), alprazolam (ALPR), amphetamine (AMP), barbiturate (BARB), buprenorphine (BUPR), cannabinoids (CANB), clonazepam (CLON), cocaine (COC), codeine (COD), diazepam (DIAZ), fentanyl (FENT), heroin (HER), hydrocodone (HCOD), hydromorphone (HMOR), lorazepam (LORA), methamphetamine (MAMP), methadone (METH), methylphenidate (MPH), morphine (MORP), oxycodone (OCOD), oxymorphone (OXYM), temazepam (TEMA), and tramadol (TRAM). When confirmatory testing was not performed, interpretations could mention drug categories [opioids (OPIO), benzodiazepines (BENZ), or amphetamines (AMP[I], distinct from amphetamine [AMP])] instead of a specific chemical structure. For each substance/category clinical interpretations could mention use (positive), no use (negative), or distance use (intermittent or low dose use).

### LLM feature extraction

To allow for predictive model training, clinical sign-out notes were first converted to substance use labels through use of an LLM model. Each interpretation note was first broken into individual sentences, and fuzzy string matching^18^ was then used to identify all sentences which mentioned any of the 23 substances or 3 categories, including non-generic names for each substances (e.g., Adderall, etc.). Each substance-containing sentence was then passed to a Bidirectional and Auto-Regression Transformer (BART) model^16^, which had been fine-tuned to provide zero-short text classifications^17^, from HuggingFace (https://huggingface.co/facebook/bart-large-mnli). The model was then queried as to whether the given sentence was ‘consistent with [positive/negative/distant use] of [substance]’. For each substance, the use pattern was returned as the label with the overall highest probability for that substance across all sentences in the interpretation. An example of this process is given in **Supplemental Fig. 1**.

Labelling accuracy was calculated by comparing outputs to human labels, across 520 random UWM interpretations from between 2022-2024. Each interpretation was manually labelled by a clinical chemist as positive, negative or distant use for each of the 26 substances/categories (13,520 labels total). All disagreements between the LLM and human expert were manually re-examined, and classified as either resulting from an LLM error, human error, or being indeterminate.

### Substance use prediction

Substance use labels were used to train ML classification models. Qualitative immunoassay results were mapped to 1 (positive) or 0 (negative), and quantitative results above or below the reporting limit were mapped to that limit (e.g., >500 to 500). Missing results were mapped to -1, and indeterminate results (e.g., due to assay interference) were mapped to -2.

Test results also included a list of what substances the ordering provider expected would be detected (based on patient disclosures, medication histories, etc.). These were mapped to a binary vector reflecting the 26 substances/categories. This data was used to train a series of gradient-boosted tree^19^ (XGBoost) models, to predict substance use patterns. Individual models were trained for each substance (n: 23), with 7 additional models trained for prediction when mass spectrometry testing was not performed. These 7 models were chosen to reflect the substances/categories that could be predicted using the immunoassay, and had sufficiently high rates of resulting without confirmatory testing: alcohol [ALC(I)], amphetamines [AMP(I)], barbiturates [BARB(I)], benzodiazepines [BENZ(I)], cannabinoids [CANB(I)], cocaine [COC(I)], and opioids [OPIO(I)]. Models were trained using an 80%-20% train-test split at the patient level, with hyper-parameter tuning performed using a random grid search and 5 cross-fold validation. Temporal stability was assessed by training models using data from 2014-2020, 2014-2021, and 2014-2022, and testing the model using all data from new patients in 2021, 2022, and 2023 respectively. All models were trained using the *scikit-learn* package in *Python 3*.*12*.

### Interpretation generation

A flow-chart process was used to convert substance use predictions into a textual interpretation. Each prediction was first tagged with metadata about the case and substance (e.g., whether the substance was a therapy, etc.), using the YAML mark-up language^20^. Substances with the same labels and tags were then grouped together (e.g., all positive therapies, all expected negative substances, etc.) and mapped to interpretative text snippets (e.g., *these findings are consistent with clonazepam and oxycodone therapy*, etc.). The snippets were then joined together to create an overall test interpretation. The text snippets were designed iteratively in consultation with the clinical chemistry team to match the writing style they typically used for sign-outs.

### Web application integration and evaluative studies

To allow for pre-deployment testing, we designed a development version of the pre-existing UWM toxicology sign-out web application^14^, that included an option to generate the AI interpretation via a single button. Clicking this button generated an interpretation and pasted it into the sign-out box, while allowing for additional editing by the user. To allow for rapid functionality, AI interpretations were pre-generated and cached for each case any time new test results became available in the application.

Alongside the AI interpretations, the dev app also integrated a new non-AI feature: a button that automatically imported a list of each patient’s recent medication dispenses into a medication list box in the app. This feature was created to improve operational efficiency, and acted as a control comparator for the integration and uptake of AI and non-AI features. To evaluate the use of both tools, 300 UDT cases from 2023 were imported into the dev app. Each case was uniformly randomized to provide access to either tool (or both, or neither). Three readers then performed a fake sign-out approximately 80 of these cases each (255 total). For each case, the sign-out time was measured, and accuracy of the AI interpretation was manually evaluated compared to the reader sign-out. Expert adjustments to the AI interpretation were classified as grammatical (e.g., minor text changes), rewording (significant text changes that do not alter meaning), adding context (additional text that provides important context but does not alter substance use patterns), or overriding an incorrect substance use statement.

### Deployment and post-deployment evaluation

The AI interpreter was deployed into live production in Aug-2025, under Clinical Laboratory Improvement Amendments (CLIA) and College of American Pathology (CAP) regulatory guidelines for custom software and laboratory information system tools, and after creation of a deployment and ongoing verification plan. All cases signed out between Aug-5-2025 and Sep-19-2025 were manually reviewed to assess whether the AI interpreter was used, and the accuracy of its predicted interpretation.

## Results

The cohort consisted of 83,553 cases, from 26,459 patients: 46.9% male, mean(std) age of 47.5(16.7)y at age of first test. Patients had a median of 1 test each, with 14.2% of patients having 5+ tests, and 4.3% having 10+ tests.

A schematic of the study design is given in **Fig. 1a-b**. The UWM urine drug test clinical workflow is illustrated in **Fig. 1c**, with estimated substance use rates given in **Fig. 1d**.

**Figure 1:**
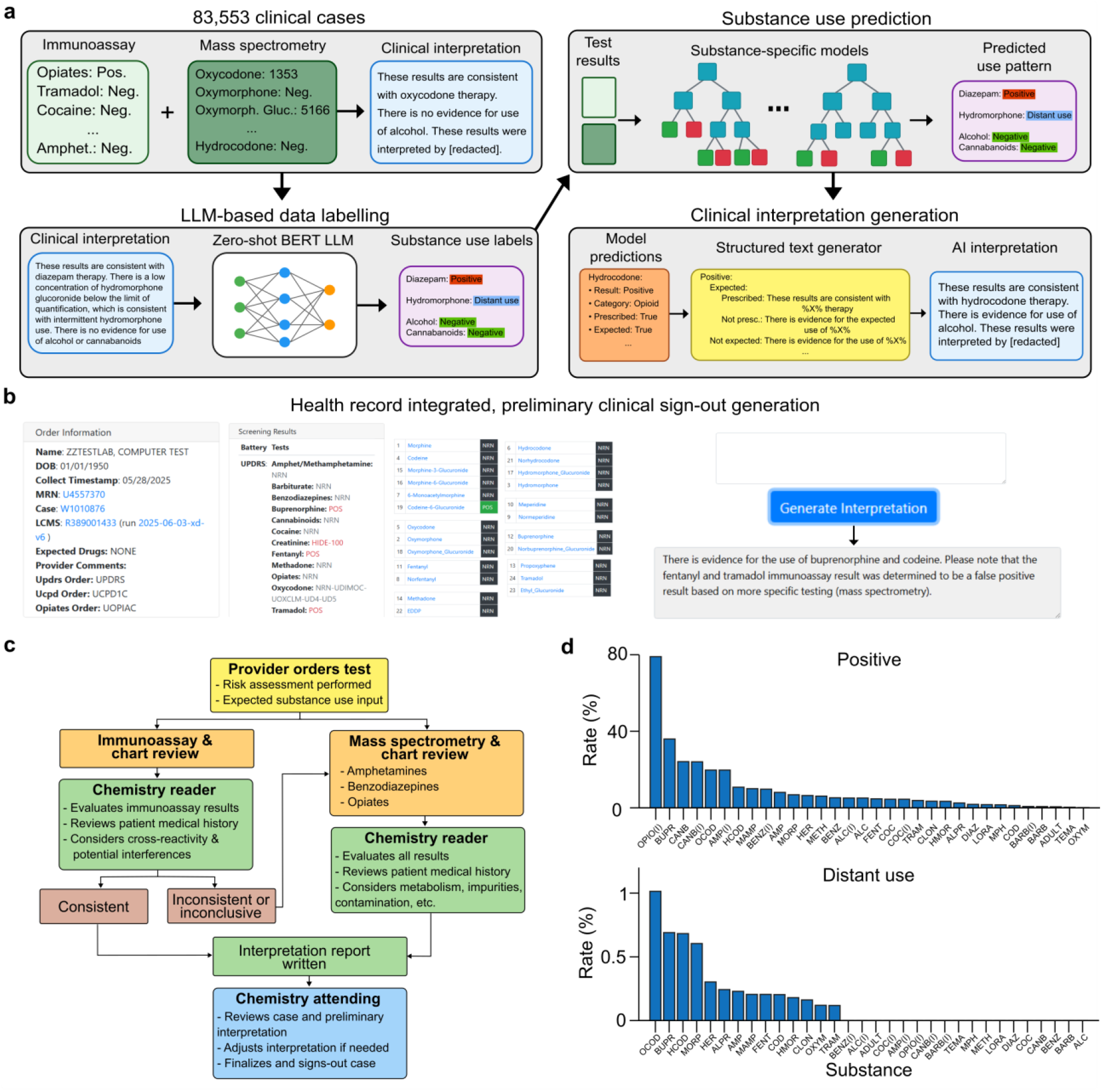
Study design and workflow overview. **a**, Overview of the study design: Large-language models were used to extract labels from historic urine drug test (UDT) notes, and used to train a substance use classification model, and create preliminary UDT sign-outs. **b**, These AI interpretations were integrated into a UDT sign-out dashboard, to allow for user testing and clinical deployment. **c**, Overview of UDT workflow at the University of Washington Medicine. **d**, Distribution of substance use patterns across the cohort. In **d**, (I) refers to classifications that were made without any confirmatory mass spectrometry testing. (e.g., BENZ refers to use of benzodiazepines [without specifying which] after mass spectrometry, and BENZ(I) refers to use of benzodiazepines where only immunoassay testing was performed). Note that results of **d**, reflect rates of use extracted from urine test interpretations, and should not be considered reflective of overall substance use rates in the UWM cohort.

### LLM labels of substance use outperform human data labelling

Compared to expert labels, LLM-based labeling of substance use patterns was highly accurate. Across 520 interpretations, the LLM achieved significantly lower error than the human labeler per substance (0.08% vs. 0.17%, p=0.05), and per case (2.1% vs. 4.2%, p=0.05) (**Fig. 2a**). LLM errors were mostly associated with high complexity cases, or on adjudications between positive and distant use of a substance. Conversely, human errors were most often in simple cases that were obvious under re-review, and so likely reflect fatigue.

**Figure 2:**
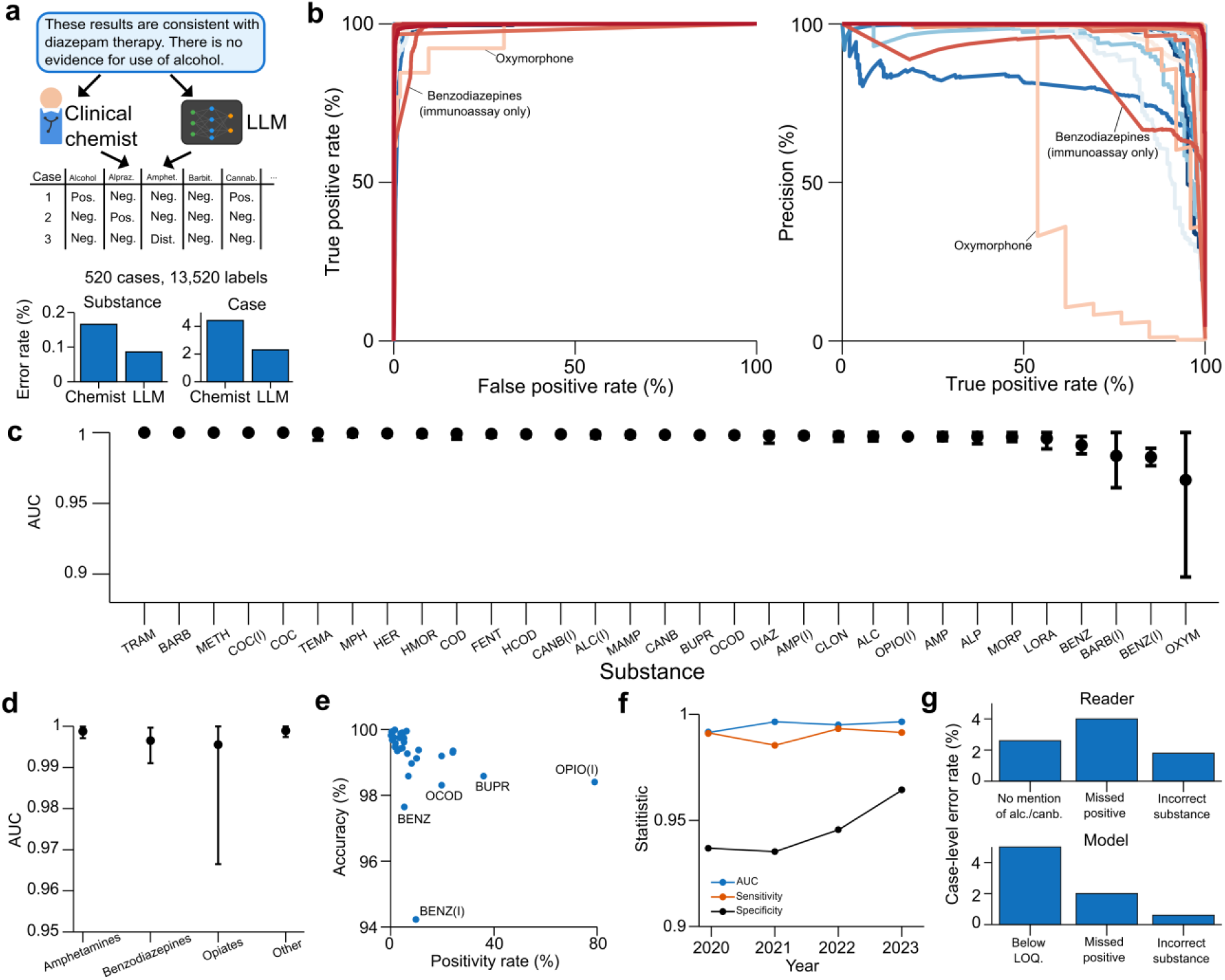
Performance of a substance use prediction model. **a**, Comparison of accuracy for manual and LLM-based labelling of substance use patterns from clinical interpretations. **b**, Receiver-operator curves for prediction of each substance use category in a held-out test set. **c**, Area-under receiver-operator characteristics (AUCs) for substance use category. **d**, Mean, min, and max AUC for each class of substances (excluding non-specific category prediction). **e**, Overall predictive accuracy against substance positivity rate. **f**, AUC, sensitivity, and specificity when training only using cases prior to a given year, and testing using that year’s data. **g**, Error rates of AI case interpretations compared to error rates of the chemistry reader, compared to the final sign-out. Error bars in **c**, reflect 95% confidence intervals generated via bootstrapping with 10,000 runs.

### AI predictions are extremely accurate for all substances

AI prediction of substance use patterns was highly accurate across all categories, with average AUC values above 0.99 (**Fig. 2c-d**), and only 3 substances falling below this threshold. These three models were for BENZ[I] and BARB[I] (which are expected to be less accurate given the lack of confirmatory testing), and OXYM (which has low prevalence [<0.2%], and which is a metabolite of oxycodone, making identification harder). Accuracy was above 94% for all substances, and above 98% for all but 2 substances (**Fig. 2e**). Performance was stable when training each model using only historic data from prior years (**Fig. 2f**).

### AI clinical interpretations are accurate and suitable

AI textual interpretations were also highly accurate. Across 161 cases, AI interpretations had similar error rates to those of the expert readers (**Fig. 2g**) (8.1% vs. 7.45% p=0.84). However, the primary reason for errors differed substantially between the two groups. Reader mistakes were most often due to not mentioning a positive substance or failing to mention use of a recreational substance such as alcohol or cannabinoids (in cases where this was then mentioned by the attending). Conversely, AI mistakes were most often due to calling a substance negative instead of distant use, due to not having access to data that was below the mass spectrometry reporting limit but above the limit of detection (which readers and attendings had manual access to).

### AI interpretations enabled faster clinical sign-outs

Under randomization of access in a replica of the clinical workflow (**Fig. 3a**), use of the AI interpreter led to significant efficiency gains. Clinicians using neither the AI tool, or a control non-AI assistive tool, averaged 126s per case sign-out. Use of the AI tool alone decreased this time by 28.5s (23%), while use of both the AI and non-AI tool decreased this time by 65s (51%) (**Fig. 3b-c**). These improvements scaled with experience, with improvements of 35%, 45%, and 65% from the most to least experienced reader.

**Figure 3:**
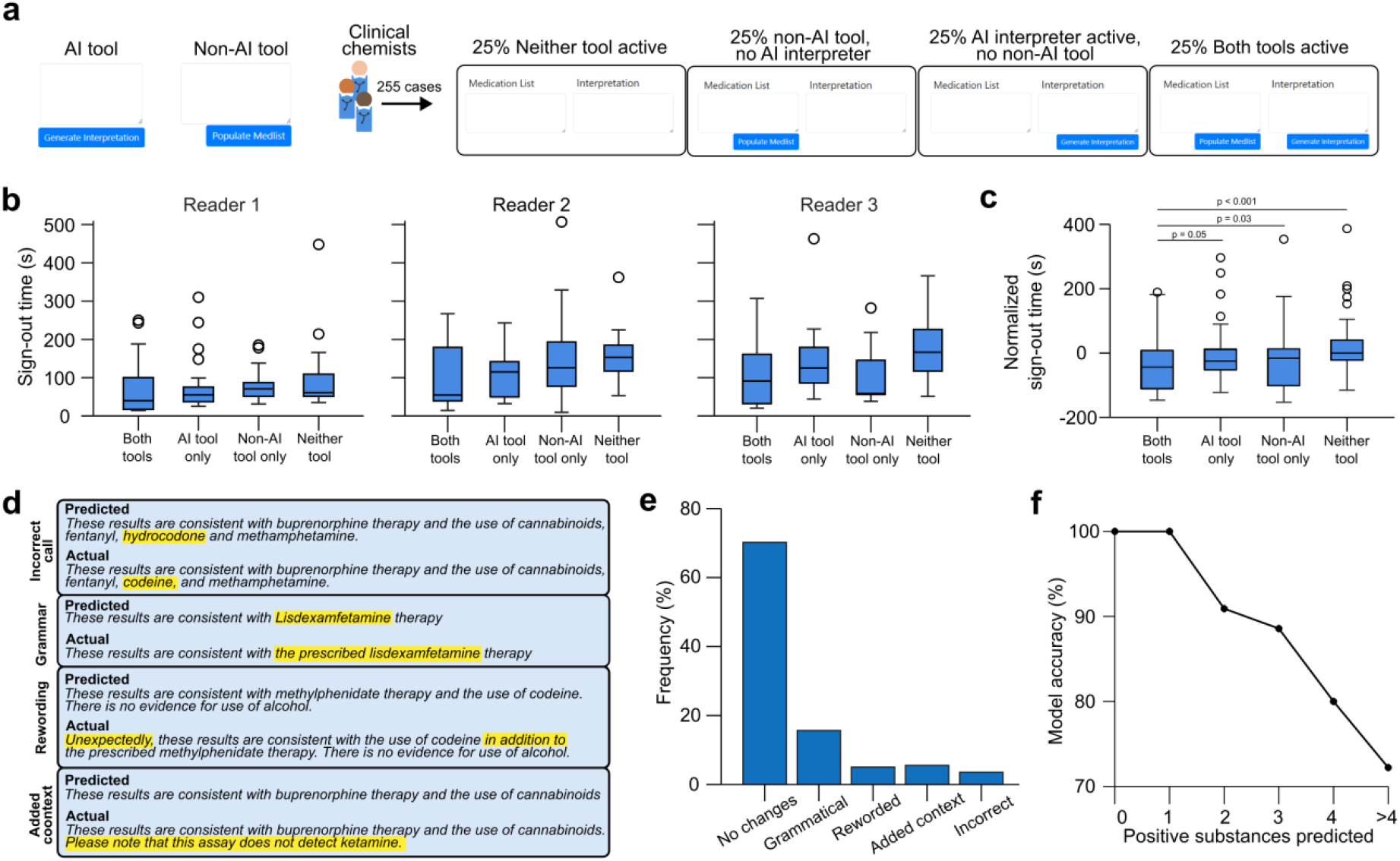
Performance of AI interpretations within the clinical workflow. **a**, Schematic of a randomization study for use of the AI interpreter alongside a non-AI workflow improvement tool. **b**, Distribution of case sign-out times for three chemistry readers when using these tools versus not. **c**, Pooled distribution of sign-out times, after normalizing each reader by their median time when using neither tool. **d**, Examples of the four types of changes the user made to AI interpretations. **e**, Frequency of reader changes to AI interpreter. **f**, Likelihood the AI interpreter will correctly predict the overall substance use pattern as the total number of predicted used substances increases.

These efficiency improvements were seen without compromising accuracy. AI interpretations were unchanged by the reader in 70% of sign-outs and only underwent light grammar modifications in a further 15% of cases (**Fig. 3d-e**). AI interpretations were only overridden due to incorrect substance use classification in 5% of cases, 40% of which were due to complex examples of sample adulteration. Accuracy of the predicted substance use pattern was highest for cases with simple substance use patterns but decreased as the total number of detected substances grew (**Fig. 3f**).

### AI interpretations retained high clinical use post deployment

After deployment, AI interpretations retained high use, being used for >70% of cases by three of the four readers on service (**Fig. 4a-b**). Usage was low (20%) for one reader, but similarly low usage was also seen for the non-AI tool, suggesting they may be resistance to workflow changes, whether AI-enabled or not. Readers were seen to modulate their use of both the AI and non-AI tools depending on which attending’s case they were evaluating, with one reader modulating their use of AI from 20% to 100% across attendings (**Fig. 4c**). Despite this, the interpretation accuracy was similar regardless of whether the reader used AI or not, and compared to what AI alone would have produced (**Fig. 4d**).

**Figure 4:**
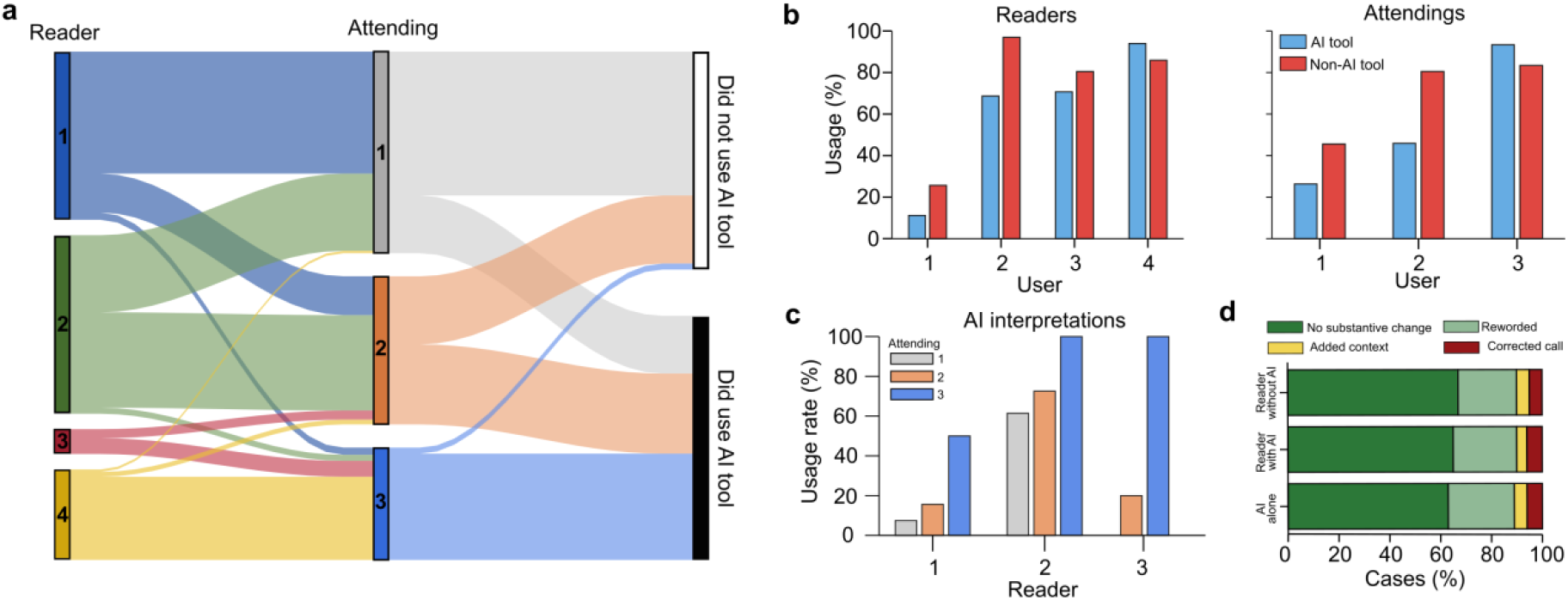
Post deployment use of AI sign-outs. **a**, Alluvial plot showing which readers and attendings reviewed each case, and whether the AI interpreter was used. **b**, Use rates of AI interpretations and non-AI tool for each reader by which attending was assigned to the case. **c**, Accuracy of preliminary sign-out when AI was or wasn’t used by the reader, compared to what AI alone would have generated. **d**, Rate that attendings reword the preliminary sign-out, based on whether AI was or wasn’t used. In **d**, AI alone refers to a comparison of the attending sign-out to what the AI interpreter would have produced if used – as no cases were signed out without a reader.

## Discussion

Here, we outline the development, deployment, and evaluation of an AI tool for generating clinical interpretations of UDTs. We show that this tool provides accurate test interpretations, and greatly improved sign-out efficiency. This is a novel example of joint use of LLMs and classical ML to enhance clinical practice.

Recent improvements in LLM capabilities have generated substantial excitement about their potential value in medicine^7^. However, few LLM tools have been clinically deployed, due to their potential for harm through factors such as model instability or hallucation^10–12^. Our study illustrates an alternate way for LLMs to support clinical operations, while minimizing these risks. Most patient health record information is stored in textual clinical notes, which are inherently harder to train predictive models with. Our findings highlight how LLM tools can now unlock this information by converting textual features into quantitative training targets, which can enable deployment of more stable classical ML models, which face significantly lower regulatory barriers than generative models^21^.

UDT testing reflects a common clinical challenge – where an ordered test generates complex data that may require specialist interpretation or consultation. This introduces substantial costs, increases test turnaround times, and may limit the testing volume a medical center can feasibly provide. In this context, our results suggest that human-in-the-loop AI test interpretation can provide significant efficiency gains, allowing centers to both save costs and offer testing to more patients. Results in **Fig. 3f** also suggest these tools may also provide triage value – enabling rapid interpretation in simple settings and allowing clinicians to focus more effort on the highest need cases. While our study focused on UDTs, we envision similar approaches being beneficial for services with similar data interpretation challenges, such as hemoglobin electrophoresis, tissue pathology analysis, and electrocardiograms.

Results in **Fig. 4** showed high usage of the AI tool post deployment, but this varied substantially across attendings. Some attendings were optimistic, and eager to integrate AI tools into their workflows, while others were more change resistant. This is consistent with other studies in the literature, which show varied clinician perspectives on the value of AI to medicine^12,22–24^. Interestingly, **Fig. 4b** showed that readers modulated their use of AI tools to align with the desires of the case’s attending. This suggests that for successful AI deployment, there is need to not just consider the users desires, but also the desires of downstream clinical stakeholders^25^. Successful deployment of AI will require not just user training, but training across the broader clinical ecosystem the tool exists within.

Our results reflect a highly novel deployment of AI for assistive sign-out of toxicology testing. AI interpretations achieved clinician level accuracy, while significantly improving sign-out efficiency. We believe this provides a strong example of how AI tools can enhance the modern clinical laboratory.

## Acknowledgements

We thank the entire UWMC clinical chemistry team for their assistance and input throughout all project stages. We thank Nathan Breit for his support in deployment and ongoing maintenance of the tool.

## Data availability

Due to restrictions on sharing protected health information, patient level data cannot be shared.

## Author contributions

BHF and PCM conceived of the study. Data collection was performed by BHF, NL, NB, and AP. Model design and training were performed by NL and BHF. Data labelling and user testing were performed by MK, HL, BA, MA, WP, AH, and GB. All authors contributed to data interpretation, manuscript writing, and editing.

